# Acetylsalicylic Acid and Ibuprofen as Adjunctive Therapy for Tuberculosis

**DOI:** 10.64898/2025.12.01.25341191

**Authors:** Lilibeth Arias, Ziyaad Waja, Nestani Tukvadze, Tumelo Moloantoa, Kennedy Otwombe, Tamta Korinteli, Judith Farrés, Chiara Sopegno, Nino Gogichadze, Kaori L. Fonseca, Natasha Pillay, Thabiso Seiphetlo, Pere-Joan Cardona, Lídia Carabias, Adrián Siles, Nadia Llavero, Carles Quiñones, Helen McShane, Willem Hanekom, Anne Ma Dyrhol-Riise, Petros C. Karakousis, Salome Charalambous, Sebastián Videla, Sergo Vashakidze, Neil Martinson, Cristina Vilaplana

**Author notes:** Shared first co-authorship (Lilibeth Arias, Ziyaad Waja, Nestani Tukvadze, Tumelo Moloantoa). Co-last authors (Sergo Vashakidze, Neil Martinson, Cristina Vilaplana). Corresponding author: Cristina Vilaplana, MD, PhD, Principal Investigator, Tuberculosis Experimental Unit (UTE) Fundació Institut Germans Trias i Pujol (IGTP), Campus Can Ruti. Crtra. De Can Ruti, Camí de les Escoles, s/n. Badalona, Barcelona, 08916, Catalonia (Spain).

## Abstract

**Background:** As novel strategies are needed to improve tuberculosis (TB) outcomes and shorten treatment; we evaluated whether host-directed therapy with non-steroidal anti-inflammatory drugs (NSAIDs) could enhance treatment response when added to standard therapy.

**Methods:** We conducted a phase 2b, double-blind, placebo-controlled, randomized trial in South Africa and Georgia. Adults with microbiologically confirmed pulmonary TB were randomized to receive standard therapy plus acetylsalicylic acid (ASA) (300 mg), ibuprofen (IBU) (400 mg), or placebo twice daily for 4 weeks and once daily for 4 weeks. Primary end points were time to a 67% reduction in the clinical symptom/sign-based TB score (TBS) and time to stable sputum culture conversion (SCC). Safety and clinical, radiological and microbiological assessments were performed up to 24 weeks after treatment completion.

**Results:** Of 221 randomized participants, 204 were included in the analysis. No significant differences were observed in the primary end points; median time to 67% TBS reduction was 4.5 weeks (ASA) and 5.0 weeks (IBU and placebo), and median time to stable SCC was 8 weeks in all groups. Both NSAIDs were associated with earlier cavity resolution at week 8, sustained at week 24 for ASA. IBU was associated with greater clinical improvement in multidrug-resistant-TB. Adverse event rates were similar across arms.

**Conclusions:** Adjunctive ASA and IBU did not shorten the time to clinical or microbiologic response but were associated with earlier cavity resolution and improved TBS in specific subgroups, with similar safety to standard therapy alone.

Funded by the European Union’s Horizon 2020; ClinicalTrials.gov number, NCT04575519.

---

Tuberculosis (TB) affects over 10 million people and causes more than 1 million deaths annually. ^1^ Up to half of survivors develop temporary or permanent respiratory disability.^2^ Novel therapeutic strategies are urgently needed to both enhance outcomes and reduce the duration of TB treatment.^3,4^ Over the past decade, a variety of host-directed therapy (HDT) candidates have been proposed as adjuncts to standard TB regimens,^5^ but clinical use is limited by cost, regulatory barriers, and lack of commercial incentives.^6^ Patient variability further complicates implementation, as benefits are not uniform.^7^ In this context, HDT with anti-inflammatory effects may improve prognosis by reducing tissue damage and the risk of permanent sequelae linked to the lung destruction and worsened clinical outcomes due to persistent inflammation in established disease.^3,7,8^

Anti-inflammatory agents such as indomethacin have historically been used empirically in inflammatory TB,^9^ corticosteroids remain standard for TB meningitis, and non-steroidal anti-inflammatory drugs (NSAIDs) are commonly used to manage early treatment symptoms.^10^ However, the use of anti-inflammatory agents in TB is largely based on clinical experience, with few systematic evaluations^11,12^ and guidelines offer limited recommendations.

Here, we report the results of the SMA-TB randomized controlled trial, conducted under the European Commission-funded SMA-TB project,^13^ which assessed the efficacy and safety of acetylsalicylic acid (ASA) and ibuprofen (IBU) as affordable, well-tolerated NSAIDs alongside standard-of-care treatment for patients with microbiologically-confirmed pulmonary TB.

## Methods

### Trial design and oversight

We conducted a phase 2b randomized, double-blind, placebo-controlled trial to estimate the potential efficacy and safety of two repurposed drugs, ASA and IBU, as adjunct add-on therapies and compared these with the standard WHO-recommended TB regimen. The trial was conducted at three clinical sites in South Africa and Georgia. Detailed trial methods and design have been previously described^14^ and are available in the **Supplementary Appendix**.

The study protocol, informed consent forms, and investigator brochures were submitted and approved by national regulatory bodies and local and institutional ethics committees. Site monitoring was conducted according to a written monitoring plan, and a data safety monitoring board was established prior to the start of the study and provided oversight throughout the trial (**Supplementary Appendix Section 1**). All participants provided written informed consent. The funders had no role in the design or conduct of the trial or in the dissemination of the data. All the authors vouch for the completeness and accuracy of the data and for the fidelity of the trial to the protocol.

### Participants

Newly diagnosed individuals with drug-susceptible (DS)-TB and multidrug-resistant (MDR)-TB were identified and once informed consent was obtained, eligibility was assessed to include adults aged 18–60 years with laboratory-confirmed pulmonary TB, irrespective of sex or HIV status.

Exclusion criteria included major comorbid conditions either requiring or contraindicating the use of NSAIDs or anticoagulants, institutionalization, TB treatment for >4 days within the past 6 months, pregnancy or breastfeeding, relevant laboratory abnormalities, weight ≤45 kg, people living with HIV (PLWH) with CD4<100 cells/mm^3^, viral load >400 copies/ml or candidates of receiving antiretroviral therapy (ART) during the first 8 weeks of TB treatment, excessive alcohol intake, or other factors that could compromise protocol adherence or the interpretation of results. Detailed inclusion and exclusion criteria are provided in the **Supplementary Appendix (Section 4).**

### Randomization and treatment

Participants were randomly assigned (1:1:1) to receive standard TB therapy plus ASA (300 mg), IBU (400 mg), or placebo, using a computer-generated sequence with block randomization stratified by site and TB resistance. Dosing was twice daily for 4 weeks, then once daily for 4 more weeks. Standard care followed National Tuberculosis Program guidelines (full details in the **Supplementary Appendix, Sections 5 and 6**).

### Procedures and follow-up

All protocol-specific procedures by timepoint are described in the **Supplementary Appendix, Section 8 and Table S2,** and included clinical examinations, adherence monitoring, safety evaluations, sputum culture conversion (SCC), radiologic evaluations, administration of health-related quality of life (HRQoL) questionnaires; and collection of samples for host and pathogen biomarker studies. Participants were followed through completion of TB treatment, with visits scheduled weekly until 2 weeks after the administration of the intervention (NSAIDs or placebo) and subsequently at weeks 12, 16, 20 and 24. An additional visit was performed at the end of TB treatment in MDR-TB participants, and nearly all participants underwent a further assessment 6 months after treatment completion.

### End points

The co-primary efficacy end points of the trial were the time to achieve a sustained reduction of at least 67% in the clinical symptom/sign-based TB score (TBS)^15,16^ over the course of TB treatment and the time to stable SCC defined by obtaining 2 consecutive negative cultures at least 4 weeks apart.

As secondary end points, we assessed: (1) differences between the intervention and placebo arms in the time to stable SCC at weeks 8 and 16 after treatment initiation; and, at baseline, week 8, and week 24, improvement or resolution of (2) clinical signs and symptoms (TBS); (3) lung function (spirometry); (4) radiological on chest X-ray (RUTI-TB Evolution Score – RTBES^17^) and evaluation of cavity resolution; and (5) HRQoL (St. Georges Respiratory Questionnaire and Kessler-10^18,19^). Predefined thresholds for TBS, RTBES, and HRQoL improvement were used to classify participants as good or poor responders. These thresholds are publicly available^20^ and detailed in **Supplementary Section 9.3**. Exploratory end points included analyses of non-specific inflammatory laboratory parameters (**Supplementary Section 9.3**).

Safety evaluation included the proportion of participants with at least one serious adverse event (SAE) in each study arm until the end of TB treatment. The SAE rate was expressed in person-time, calculated from the day of the first dose of NSAID or placebo until 30 days after the last dose, including all adverse events (AE) recorded in each arm. Tolerability was assessed by the proportion of participants who permanently discontinued NSAID or placebo.

### Analysis population

The study population included participants with DS-TB and MDR-TB. The safety population consisted of all randomized participants who received at least one dose of the study drug. The modified intention-to-treat population included all randomized participants, excluding those who were withdrawn after baseline without any post-baseline primary end-point data or who met late exclusion criteria.

### Statistical Analysis

The sample-size calculation (**Supplementary Appendix, Section 7)** was based on DS-TB participants, comparing the standard-of-care plus placebo arm with each intervention arm under identical assumptions. An MDR-TB population was enrolled but excluded from sample-size calculations and the primary end points analysis. For sensitivity analyses, MDR-TB participants were combined with DS-TB participants within treatment arms, and efficacy was re-evaluated using the same methodology.

Baseline socio-demographic characteristics were summarized using medians and interquartile ranges for continuous variables, and frequencies were determined for categorical variables. Continuous measures were compared across groups using the Kruskal-Wallis test.

For the two primary end points, time-to-event data were compared among treatment groups with the log-rank test. Hazard ratios and 95% confidence intervals (CI) were estimated with a Cox proportional-hazards model. No adjustment for multiplicity was applied.

Secondary end points were analyzed as follows: for secondary end point 1, Cox proportional hazards regression was used to compare NSAIDs arms with the placebo arm. For end points 2-5, the evolution of the individual scores over time was compared between groups using linear mixed models, with p-values calculated for each visit and an overall global p-value. For end points 2, 4, and 5, the proportion of individuals achieving the desired improvement was compared between groups by calculating the risk difference, with corresponding 95% CI derived using Newcombe’s method. Exploratory end points were compared using the chi-square test for categorical variables.

For safety analyses, event frequencies and incidence rates per 100 person-weeks, with corresponding 95% CI, were estimated for each study group and compared using a Poisson regression model, with the placebo group as the reference. Time to first AE or SAE, determined up to 30 days after the last dose, was analyzed with the Kaplan–Meier method, and differences between the intervention and placebo groups were assessed with the log-rank test. Statistical analyses were conducted in SAS Enterprise Guide (version 8.3), Stata 15, Python (version 3.11) and R (version 4.3.2).

## Results

### Participants

The first patient was screened in February 2021, and the last patient completed the final visit in March 2024. A total of 426 newly diagnosed individuals with pulmonary TB were screened for eligibility. Among those eligible, 221 participants were randomly assigned to receive ASA, IBU, or placebo and were included in the safety population (146 in South Africa and 75 in Georgia). Of these, 204 participants met modified intention-to-treat criteria. Primary end points were analyzed in DS-TB population, while baseline characteristics and secondary end points were also analyzed in the combined DS-TB and MDR-TB population. (**Fig. 1**). The demographic and clinical characteristics of the participants were similar between the three arms with higher percentage of male participants (73%), and the median age 35 years (**Table 1** and **Table S4**).

**Figure 1:**
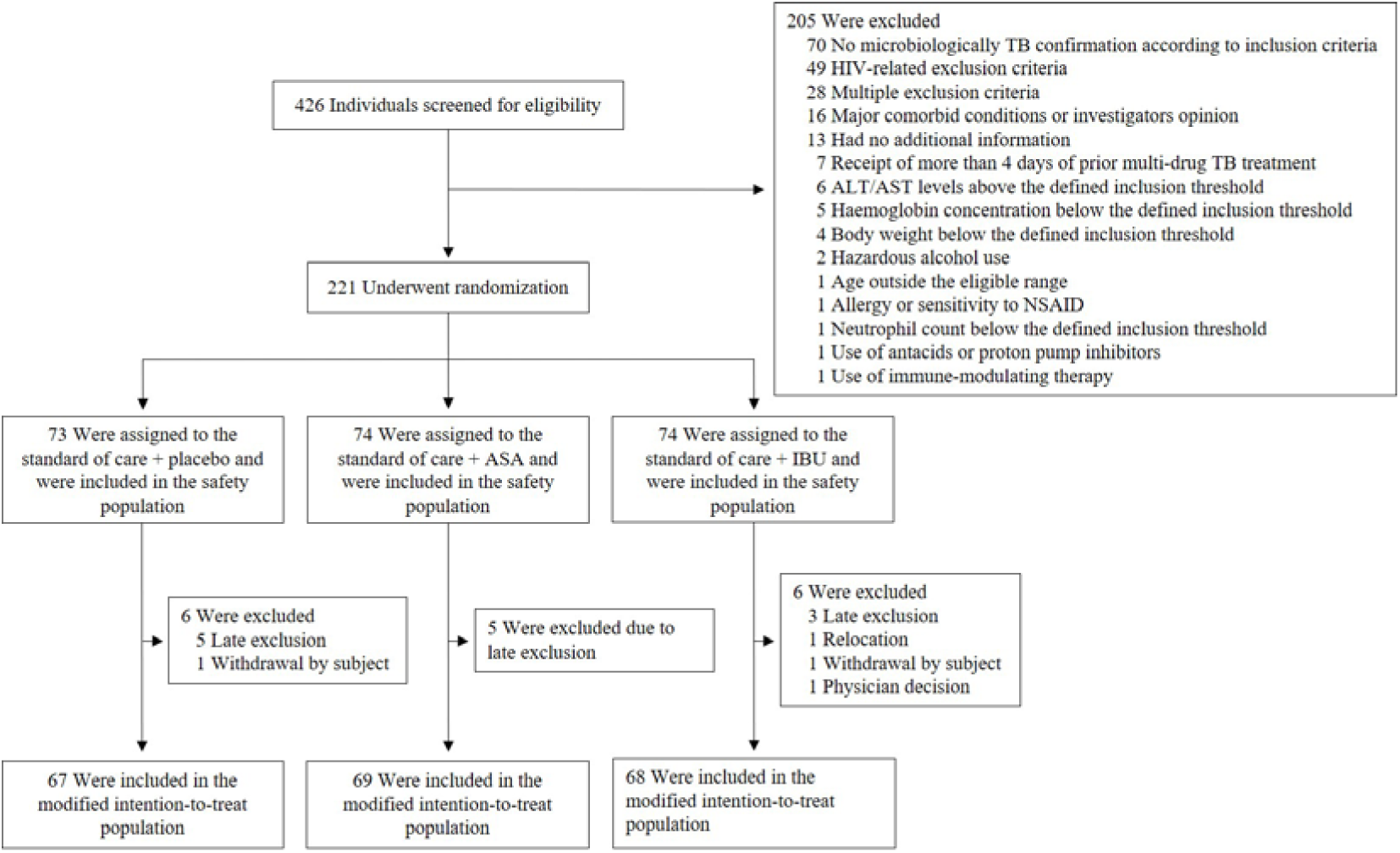
Screening, Randomization, and Analysis Populations. A total of 426 individuals were screened, and 221 underwent randomization in a 1:1:1 ratio to receive standard therapy plus placebo, acetylsalicylic acid (ASA), or ibuprofen (IBU). The most common reasons for exclusion were absence of microbiologic confirmation of TB and HIV-related criteria. A total of 204 participants were included in the modified intention-to-treat analysis.

**Table 1:**
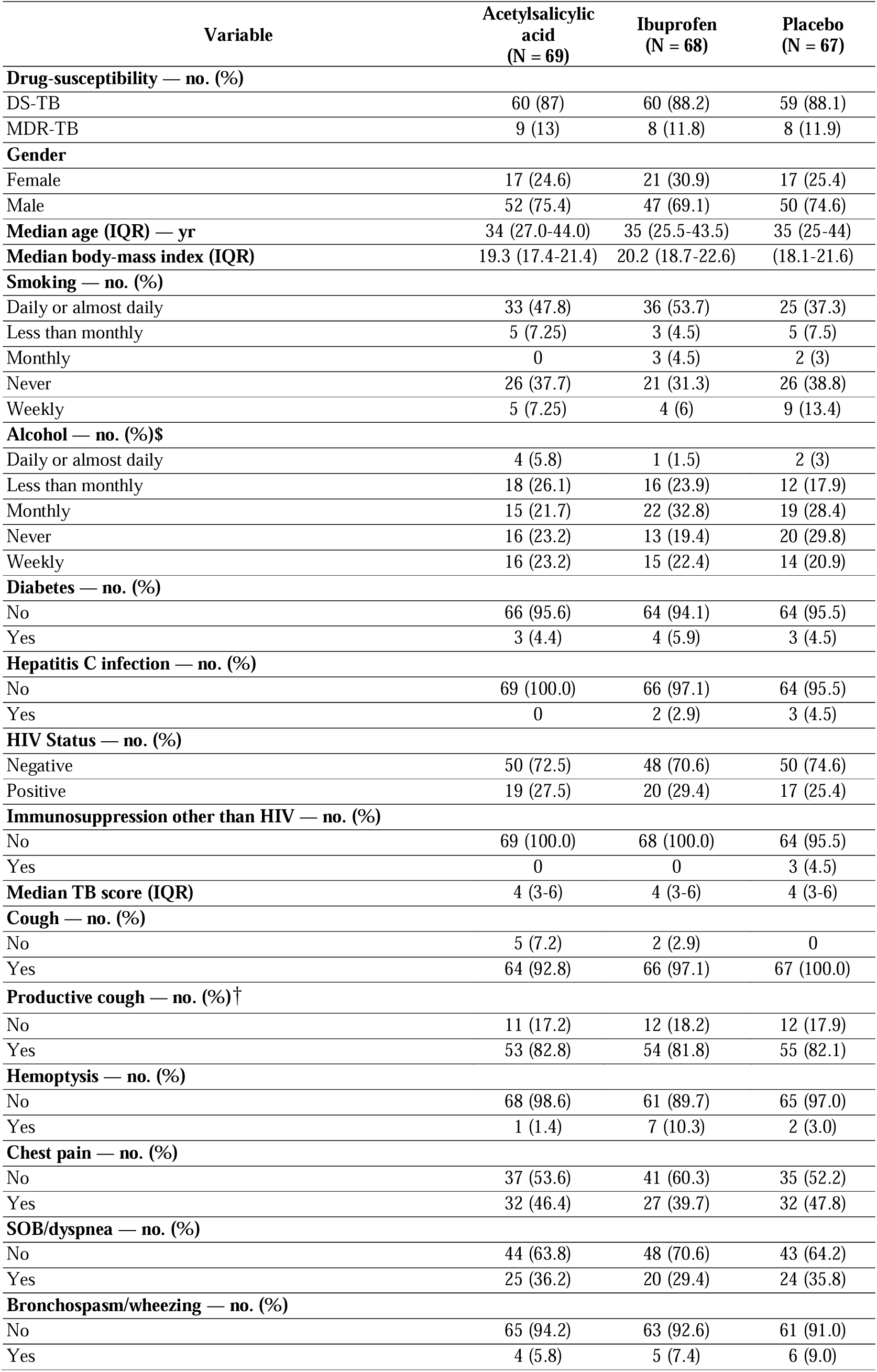
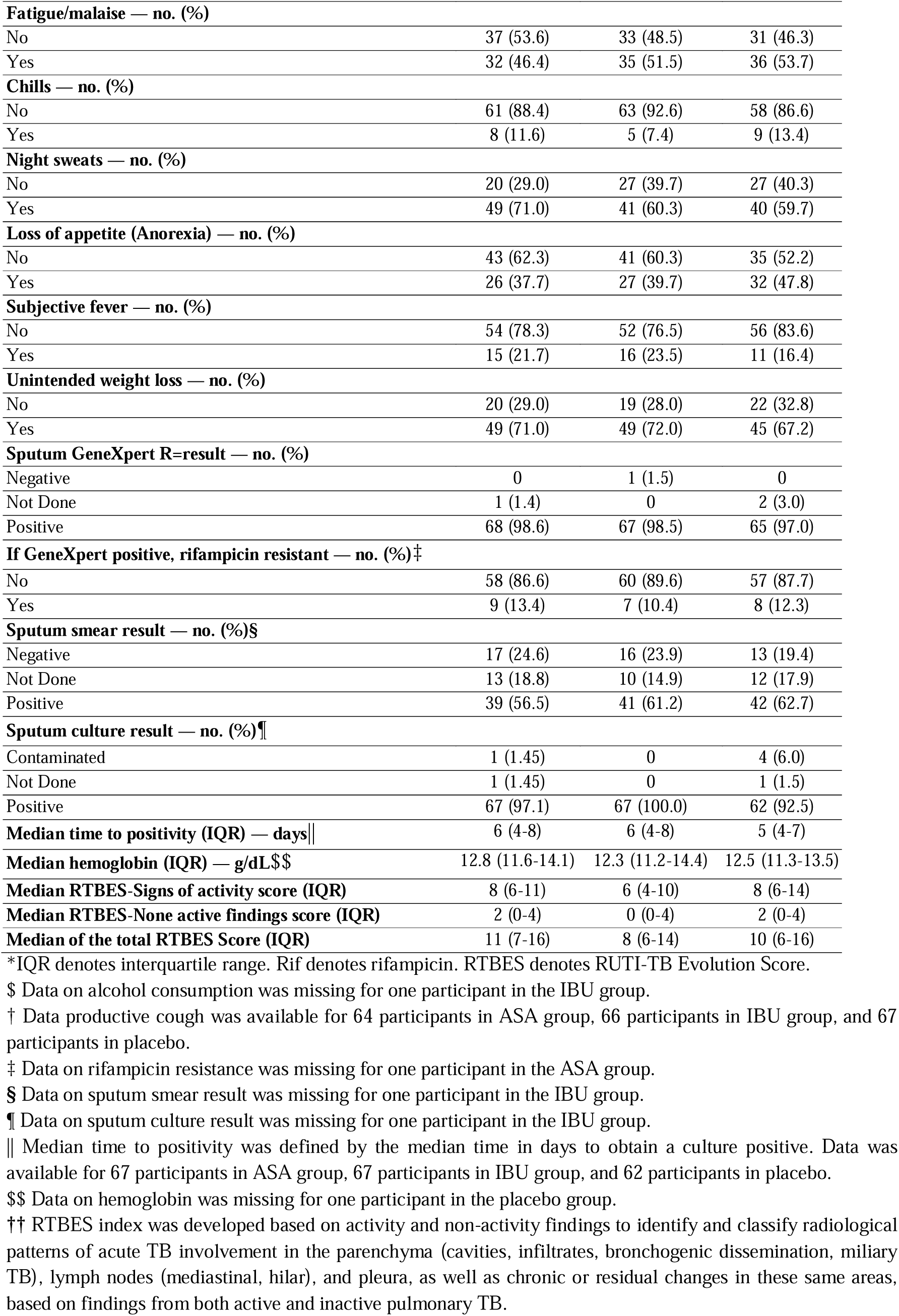
Baseline characteristics.

### Primary efficacy results

There was no statistically significant difference between arms for the primary efficacy end points when analyzing DS-TB participants. The median time to achieve a 67% reduction in TBS was 4.5 weeks (95% CI, 2.3 to 6) in ASA, 5 weeks (95% CI, 3.1 to 6.7) in IBU and 5 weeks (95% CI, 4 to 7) in placebo (**Fig. 2a and Table S5**). Likewise, the median time to stable SCC was 8 weeks (95% CI, 6 to 8) in the ASA group, 8 weeks (95% CI, 6.1 to 8) in the IBU group and 8 weeks (95% CI, 8 to 11.9) in the placebo group (**Fig. 2b and Table S5**). These results were consistent in the combined DS/MDR-TB cohort (**Table S5 and Fig. S2A and S2B)**.

**Figure 2:**
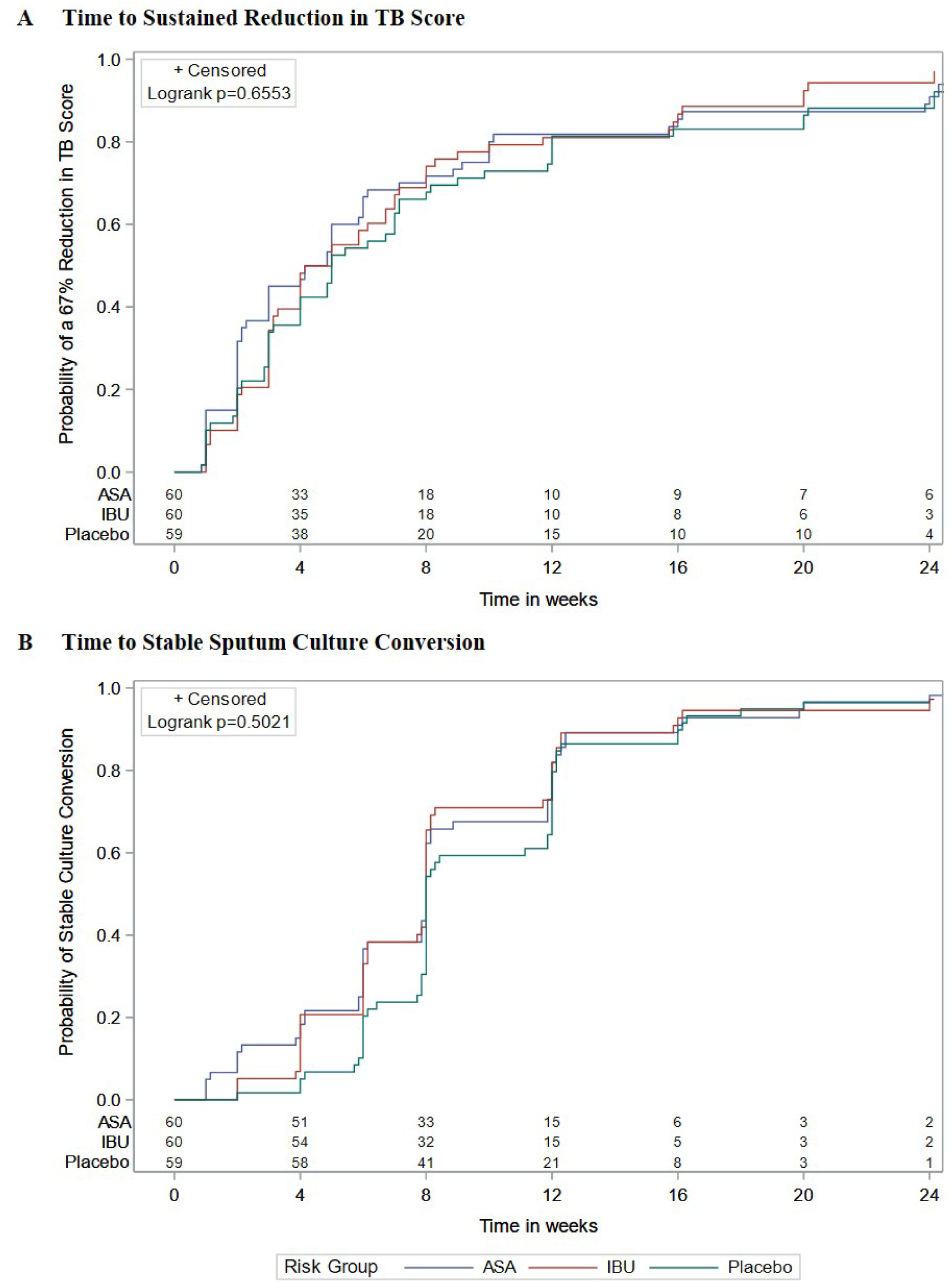
Primary efficacy end points at week 24. Figure shows Kaplan–Meier curves stratified by study arm generated to assess group differences in DS-TB participants. Panel A shows the time to a 67% reduction in TB score. Panel B shows the time to stable sputum culture conversion. ASA denotes acetylsalicylic acid and, IBU ibuprofen.

### Secondary and exploratory end points

No significant differences were observed between ASA or IBU and placebo in the proportion of participants achieving a ≥67% reduction in TBS or stable SCC at week 8 or week 16, consistent with the primary end point findings at week 24 (**Table S5 and Fig. S3 and S4**).

Participants with DS-TB who received NSAIDs were significantly more likely to achieve cavity resolution by week 8 compared to those receiving placebo, with risk differences of 0.22 (95% CI, 0.07 to 0.36) for ASA and 0.14 (95% CI, 0.00 to 0.27) for IBU (**Fig. 3**), with similar magnitudes in the combined DS-TB and MDR-TB population (**Figs. S5 and S6).** This effect persisted through week 24 for ASA only (0.25; 95% CI, 0.04 to 0.43; **Fig. 3**). Among participants with MDR-TB, IBU treatment was associated with greater of TBS improvement at both weeks 8 and 24, with risk differences of 0.50 (95% CI, 0.05 to 0.78) and 0.61 (95% CI, 0.10 to 0.82), respectively (**Fig. S6**).

**Figure 3:**
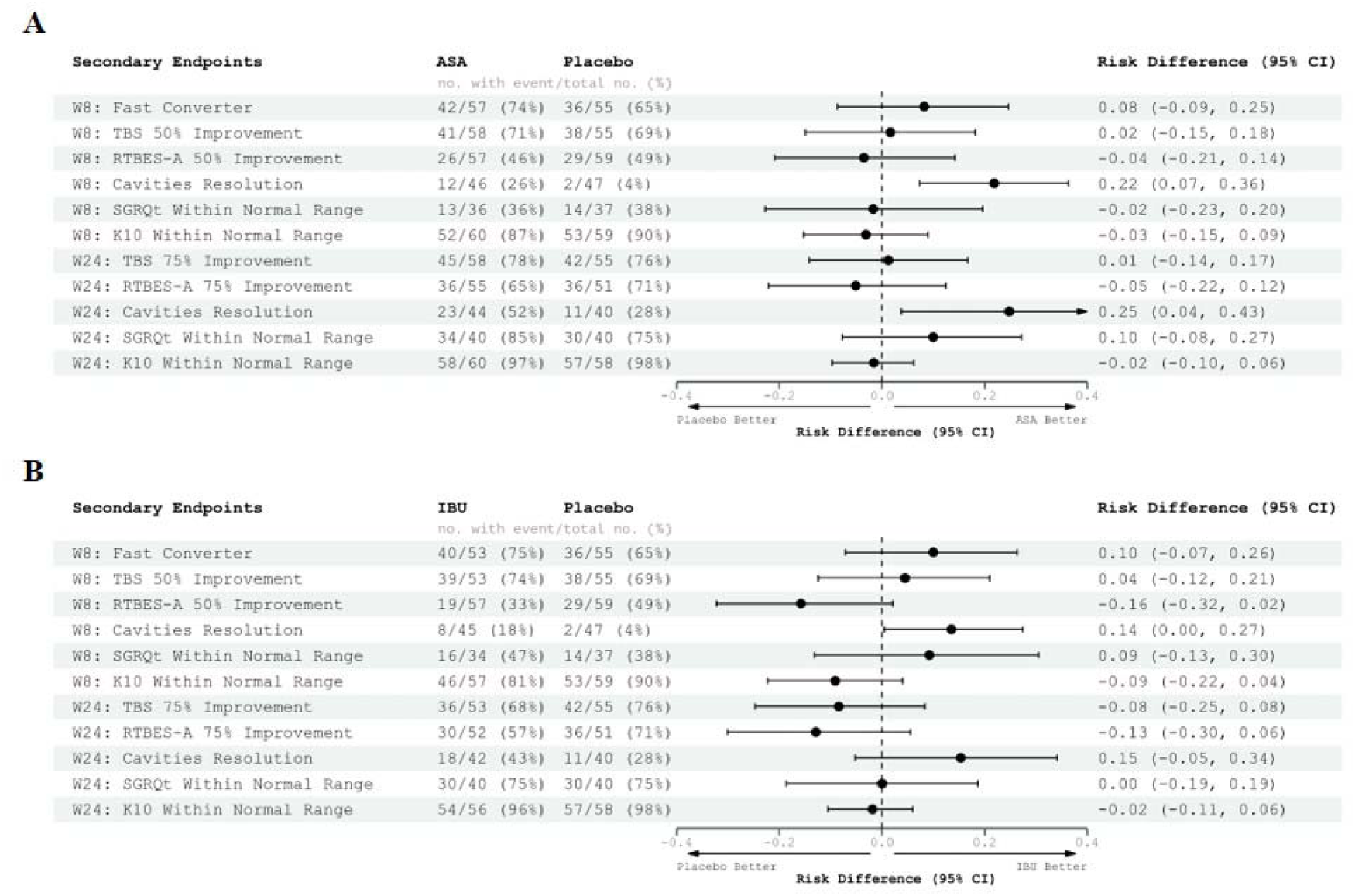
Secondary end points. A. Results for ASA arm. B. Results for IBU arm. Risk difference between NSAIDs treatment and placebo group for each of the secondary end points proposed to assess the patients’ improvement. The proportions of patients showing improvement in the two treatment groups are reported. For the risk difference a 95% confidence interval is computed using Newcombe’s method. TBS denotes TB score, RTBES-A RUTI-TB Evolution Activity Score, SGRQt Saint Georges Respiratory Questionnaire, and K10 Kessler-10 Questionnaire.

The evaluated population showed improvement in clinical, radiological and lung functional parameters throughout the treatment measured at weeks 8 and 24, with no statistically significant differences observed between either NSAID arms versus placebo (**Fig. S7 and Table S6**).

Finally, the NSAIDs arms showed a reduction in systemic inflammatory markers, including erythrocyte sedimentation rate (ESR) and monocyte-lymphocyte ratio (MLR), with values shifting toward the normal range compared to placebo significant at multiple time points, suggesting a potential immunomodulatory effect of NSAIDs (**Fig. S10**).

### Safety

Both NSAIDs were shown to be safe. A total of 93 AEs were reported in 70 participants during or up to 30 days after administration of the last dose of NSAID or placebo. The incidence of AEs per 100 person-weeks (up to 90 days) was similar to that observed with placebo (rate ratio, 1.3; 95% CI, 0.8–2.0; P=0.33 for ASA, and 0.8; 95% CI, 0.5–1.4; P=0.52 for IBU). Thirty-four AEs were classified as grade 3 or higher and 13 were classified as SAEs, with no statistically significant differences across the three study groups; none were considered definitely related to the investigational product (IP) (**Table 2**). The most common AEs (in >5% of the participants in any of the groups) were dyspepsia, toxic drug-induced hepatitis and hypertension (**Table S9**). The safety analysis for the 24-week TB treatment is provided in **Table S10.**

**Table 2:**
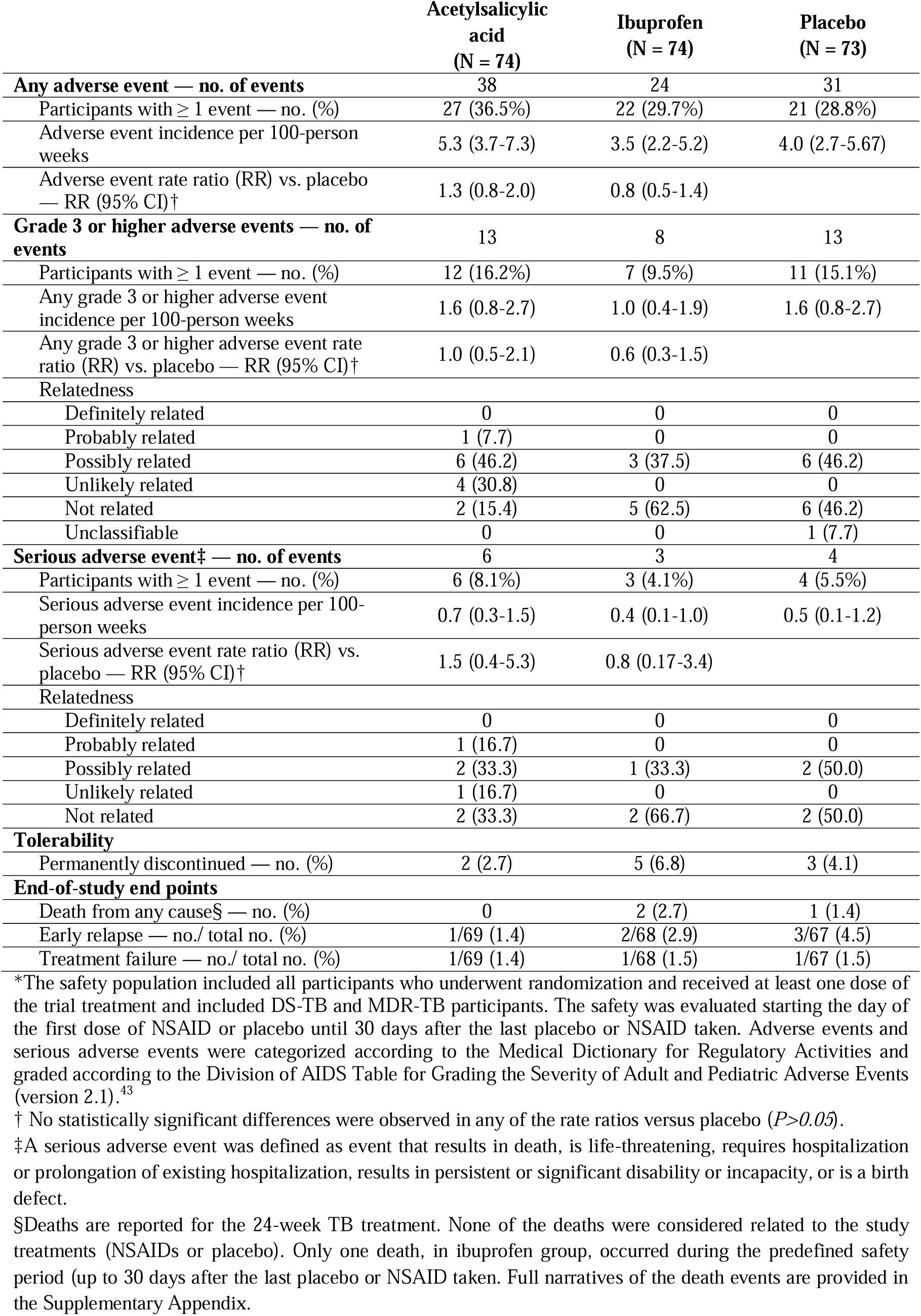
Safety results up to 90 days.

|  | Acetylsalicylic<br>acid<br>(N = 74) | Ibuprofen<br>(N = 74) | Placebo<br>(N = 73) |
| --- | --- | --- | --- |
| **Any adverse event — no. of events** | 38 | 24 | 31 |
| Participants with $\geq 1$ event — no. (%) | 27 (36.5%) | 22 (29.7%) | 21 (28.8%) |
| Adverse event incidence per 100-person weeks | 5.3 (3.7-7.3) | 3.5 (2.2-5.2) | 4.0 (2.7-5.67) |
| Adverse event rate ratio (RR) vs. placebo — RR (95% CI)<sup>†</sup> | 1.3 (0.8-2.0) | 0.8 (0.5-1.4) |  |
| **Grade 3 or higher adverse events — no. of events** | 13 | 8 | 13 |
| Participants with $\geq 1$ event — no. (%) | 12 (16.2%) | 7 (9.5%) | 11 (15.1%) |
| Any grade 3 or higher adverse event incidence per 100-person weeks | 1.6 (0.8-2.7) | 1.0 (0.4-1.9) | 1.6 (0.8-2.7) |
| Any grade 3 or higher adverse event rate ratio (RR) vs. placebo — RR (95% CI)<sup>†</sup> | 1.0 (0.5-2.1) | 0.6 (0.3-1.5) |  |
| **Relatedness** |  |  |  |
| Definitely related | 0 | 0 | 0 |
| Probably related | 1 (7.7) | 0 | 0 |
| Possibly related | 6 (46.2) | 3 (37.5) | 6 (46.2) |
| Unlikely related | 4 (30.8) | 0 | 0 |
| Not related | 2 (15.4) | 5 (62.5) | 6 (46.2) |
| Unclassifiable | 0 | 0 | 1 (7.7) |
| **Serious adverse event<sup>‡</sup> — no. of events** | 6 | 3 | 4 |
| Participants with $\geq 1$ event — no. (%) | 6 (8.1%) | 3 (4.1%) | 4 (5.5%) |
| Serious adverse event incidence per 100-person weeks | 0.7 (0.3-1.5) | 0.4 (0.1-1.0) | 0.5 (0.1-1.2) |
| Serious adverse event rate ratio (RR) vs. placebo — RR (95% CI)<sup>†</sup> | 1.5 (0.4-5.3) | 0.8 (0.17-3.4) |  |
| **Relatedness** |  |  |  |
| Definitely related | 0 | 0 | 0 |
| Probably related | 1 (16.7) | 0 | 0 |
| Possibly related | 2 (33.3) | 1 (33.3) | 2 (50.0) |
| Unlikely related | 1 (16.7) | 0 | 0 |
| Not related | 2 (33.3) | 2 (66.7) | 2 (50.0) |
| **Tolerability** |  |  |  |
| Permanently discontinued — no. (%) | 2 (2.7) | 5 (6.8) | 3 (4.1) |
| **End-of-study end points** |  |  |  |
| Death from any cause<sup>§</sup> — no. (%) | 0 | 2 (2.7) | 1 (1.4) |
| Early relapse — no./ total no. (%) | 1/69 (1.4) | 2/68 (2.9) | 3/67 (4.5) |
| Treatment failure — no./ total no. (%) | 1/69 (1.4) | 1/68 (1.5) | 1/67 (1.5) |

\*The safety population included all participants who underwent randomization and received at least one dose of the trial treatment and included DS-TB and MDR-TB participants. The safety was evaluated starting the day of the first dose of NSAID or placebo until 30 days after the last placebo or NSAID taken. Adverse events and serious adverse events were categorized according to the Medical Dictionary for Regulatory Activities and graded according to the Division of AIDS Table for Grading the Severity of Adult and Pediatric Adverse Events (version 2.1).<sup>43</sup>
<sup>†</sup> No statistically significant differences were observed in any of the rate ratios versus placebo ( $P > 0.05$ ).
<sup>‡</sup>A serious adverse event was defined as event that results in death, is life-threatening, requires hospitalization or prolongation of existing hospitalization, results in persistent or significant disability or incapacity, or is a birth defect.
<sup>§</sup>Deaths are reported for the 24-week TB treatment. None of the deaths were considered related to the study treatments (NSAIDs or placebo). Only one death, in ibuprofen group, occurred during the predefined safety period (up to 30 days after the last placebo or NSAID taken. Full narratives of the death events are provided in the Supplementary Appendix.

A total of 3 participants died during the study (two in the IBU group and one in the placebo group). None of these deaths were considered related to the IP. Across all participants, three treatment failures occurred (two in DS-TB and one in MDR-TB cases, one per treatment arm: placebo, IBU, and ASA). Among those who completed follow-up (82.8%), six early relapses were reported: three in the placebo group, two in the IBU group, and one in the ASA group (**Table 2**).

## Discussion

This trial evaluated the efficacy and safety of ASA and IBU as adjuncts to standard TB treatment, systematically assessing the common use of NSAIDs in clinical practice for TB management. ASA and IBU were selected for their potential therapeutic benefits,^8,12^ favorable safety profiles, low cost, and broad availability. No benefit was observed in the time required to achieve a 67% reduction in TBS or stable SCC. Nevertheless, both NSAIDs had meaningful effects on early cavity resolution and safety profile were similar across arms.

In this randomized clinical trial, the primary efficacy end point was the time to a 67% reduction in TBS among DS-TB patients, hypothesizing that, if beneficial, adjunctive NSAID therapy would accelerate clinical improvement over the 24-week treatment period. TBS is a validated tool for monitoring treatment response,^15,16^ and failure to achieve a 25% reduction predicts poor outcomes and mortality.^3^ However, limited data on expected relative reductions rendered our 67% theoretical threshold, and no difference was observed between the study arms.

Because the prespecified 67% reduction threshold had limited empirical support and did not differentiate groups, we evaluated additional predefined clinical and radiologic outcomes to better characterize treatment response and variability across subgroups, ^21^ revealing substantial inter-individual variability with benefits for specific end points and populations.

Our second primary end point was the time to stable SCC, an established measure in TB trials. Delayed SCC indicates poor microbiological control and has been linked to a sustained proinflammatory profile at the lesion site,^22^ so this parameter was included to standardize comparisons and facilitate benchmarking.^10,23–26^ Although NSAIDs were not anticipated to influence SCC, a faster conversion could support shorter antibiotic courses, as demonstrated by the BPaLM regimen,^27^ whereas delayed SCC might signal antagonism, as seen with aspirin and isoniazid in murine models.^28^ Although a positive trend toward earlier SCC was observed in the NSAID arms, particularly during the early weeks, no significant differences in SCC were found among the three study arms.

The key finding of this trial was that adding either NSAID to standard TB therapy accelerated early cavity resolution, with the effect persisting for 4 months after ASA discontinuation, and without any negative effect on SCC. Previous HDT trials have shown trends toward cavity size reduction or faster cavity resolution.^29,30^ The presence of pulmonary cavities has been linked to greater disease severity, delayed SCC, and poorer HRQoL during the first year after treatment.^31–33^ Thus, the accelerated cavity resolution observed in our trial may be clinically relevant, as faster cavity healing has been suggested to indicate a lower risk of post-TB lung disease beyond microbiological cure.^34^

In terms of safety, ASA and IBU are over-the-counter medicines on the WHO Essential Medicines List, and their risks are dose-dependent and primarily gastrointestinal.^5,11,35,36^ In our trial, both were well-tolerated during prolonged administration, which is particularly important given the complex toxicity profiles of many anti-TB regimens.^37^

Although follow-up in this trial was limited to 6 months after TB treatment and completed in approximately 80% of participants, six early relapses were identified, most of which occurred in the placebo arm. While the number of events was small and did not allow for formal statistical comparison, the unequal distribution suggests a possible benefit of adjunctive therapies in reducing relapse. These findings align with observational evidence that repurposed drugs may lower TB recurrence and support further investigation.^38^ Extended follow-up of individuals enrolled in past, current and future trials will be critical to fully assess the long-term impact of HDTs on relapse prevention and sustained treatment success.

In our study, systemic inflammation was associated with treatment outcomes, consistent with prior findings.^39,40^ Although NSAID administration reduced systemic inflammatory markers during treatment,^12^ this effect did not result in significant improvements across most clinical outcomes. While these findings support the underlying principle of our research, they indicate that optimizing the timing and intensity of anti-inflammatory intervention – for example, by administering higher doses over a shorter initial period, lower doses for a longer duration, or by evaluating alternative HDTs with anti-inflammatory potential – may be necessary to achieve broader clinical benefits.

The most important limitation of our study was the early termination of enrollment after accrual of 64.7% of participants because of financial and administrative constraints exacerbated by the COVID-19 pandemic.^14^ Although enrollment was incomplete, an interim analysis indicated that 300–350 participants would likely have been required to detect statistically significant differences in the primary endpoints.

Finally, our findings highlight substantial heterogeneity in response to adjunctive NSAIDs, reinforcing the need for individualized TB treatment. As shown in MDR-TB management,^41^ personalized regimens require integration of both host– and pathogen-related predictors of disease course and therapeutic response. In our trial, effects varied by compound, outcome, and subgroup. Future research should prioritize large, diverse cohorts with planned subgroup analyses or targeted trials in defined populations to clarify efficacy and safety. Both strategies are critical to advancing tailored, evidence-based TB care.^42^

In conclusion, in this phase 2 trial, NSAIDs (ASA and IBU) added on standard tuberculosis treatment did not reduce the time to achieve a sustained clinical signs/symptoms reduction, nor the time to stable sputum culture conversion. However, this trial provides evidence that adjunctive NSAIDs use during TB treatment was associated with improved cavity resolution and improvements in TB score in specific subgroups, with a safety profile similar to standard therapy alone. These findings indicate that HDT may influence selected short-term measures of response and could have implications for long-term complications, including relapse and post-TB lung disease. Further studies are needed to define optimal dosing and duration, to determine the populations most likely to benefit, and to assess whether early improvements lead to durable clinical effects.

## Acknowledgements

We thank Dr Rosa Morros and Dan Ouchi-Vernet (IDIAP Jordi Gol) for generating the randomization list and all the individuals who participated in this clinical trial. We are grateful to Professor Andrew Copas (UCL MRC CTU, London, UK) for his expert advice on the statistical analysis plan following the DSMB and TSC recommendations, and to Professor Steffen Stenger (Institute for Medical Microbiology and Hygiene, Ulm University Hospital, Germany) for his invaluable guidance and support throughout the SMA-TB project.

## Data availability

Protocol and aggregated data from the RCT are available along with this manuscript. To submit inquiries related to SMA-TB clinical research and database please contact the corresponding author. Database will remain private until publication. De-identified data from the trial database from participants who agreed to future use of data will be provided, and its use will be limited to this project or future projects with the aim to study tuberculosis disease, how to treat it or how to prevent it. Only the minimum subsets of data needed to conduct the proposal will be provided. The data will be available to individuals or research groups whose proposed use of the data receives approval from the SMA-TB data-access committee established for review of such requests; the proposal has obtained an ethical approval; that proposal avoids duplication of work, adds value, and avoids unnecessary competition; and that the protocol complies with the minimisation use of data as per EU regulations.

## Author contributions

CV conceptualized the study and obtained the funding. She developed the protocol with SV and NM and coordinated the project with LA. ZW, NT, TM, TK, NM, and SV enrolled participants and oversaw clinical follow-up and data collection at their respective sites. LA, NG, KO, ZW, NT, TM, JF, CS, and CV curated the database. KO performed the primary statistical analysis. CS, JF, KF, NG, LA, and KO conducted the analyses of the secondary outcomes. CV, SV, NT, NM, and LA supervised the project. HMcS, WH, and AD-R were members of the Trial Steering Committee, and SC and PK were members of the Data Safety Monitoring Board; all provided essential guidance to the trial. LA and CV drafted the manuscript. CV had final responsibility for the decision to submit the manuscript. All authors contributed to data interpretation, critically reviewed and revised the manuscript, and approved the decision to submit it for publication. All authors provided written feedback during manuscript development and were directly involved in the conduct of the study.

## Use of AI-Assisted Technologies

The authors used OpenAI’s ChatGPT to improve the readability of the manuscript draft. After using this tool, the authors reviewed and edited the content as needed and take full responsibility for the content of the publication.

